# Exploring perspectives from stroke survivors, carers and clinicians on virtual reality as a precursor to using telerehabilitation for spatial neglect post-stroke

**DOI:** 10.1101/2020.01.07.20016782

**Authors:** Helen Morse, Laura Biggart, Valerie Pomeroy, Stéphanie Rossit

## Abstract

Spatial neglect is a common and severe cognitive consequence of stroke, yet there is currently no effective rehabilitation tool. Virtual Reality (VR) telerehabilitation tools have the potential to provide multisensory and enjoyable neuropsychological therapies and remotely monitor adherence without the presence of a therapist at all times. Researchers and industry need to better understand end-user perspectives about these technologies to ensure these are acceptable and user-friendly and, ultimately, optimize adherence and efficacy. Therefore, this study aims to explore end-user perspectives on the use of self-administered VR for spatial neglect in a university environment to identify barriers and facilitators prior to extending its use remotely or within the home as a VR telerehabilitation tool. We used a mixed-method design including focus groups, self-administered questionnaires and individual interviews with stroke survivors (N = 7), their carers (N = 3) and stroke clinicians (N = 6). End-user perspectives identified clarity of instructions, equipment (cost, available resources) and for some, level of experience with technology as barriers of use. Perceived facilitators of use were performance feedback, engagement and enjoyment, and psychological benefits associated by self-administered VR telerehabilitation. Overall, end-users were positive and interested in using VR telerehabilitation for spatial neglect. These perspectives enabled us to produce practical recommendations to inform development, enhance engagement and uptake of self-administered VR telerehabilitation and inform feasibility and usability studies.

## 1. Introduction

Around 20-40% of the 1.2 million stroke survivors currently living in the UK (Stroke Association, 2018) are estimated to have spatial neglect (Ringman et al., 2004; Rowe et al., 2019; Puig-Pijoan et al., 2018). Spatial neglect is a severe neuropsychological syndrome generally defined as a failure to respond to stimuli on the side of the space opposite to the side of the brain lesion. The clinical impact of spatial neglect is substantial with 40% of people showing neglect symptoms even more than a year post-stroke (Nijboer, Kollen & Kwakkel, 2013) and its presence being a major predictor of disability (e.g., Gillen, Tennen & McKee, 2005). In line with this, previous research has found that stroke survivors, carers and clinicians identified visual problems as one of the top 10 priorities in stroke research (Pollock et al., 2014). Numerous rehabilitation methods have been developed for spatial neglect (Bowen, Hazelton, Pollock, & Lincoln, 2013; Azouvi, Jacquin-Courtois & Luauté, 2017; Gammeri, Iacono, Ricci & Salatino, 2020). These can be divided into bottom-up approaches (e.g., prism adaptation, eye-patching and limb activation training) and top-down approaches (e.g., visual scanning training, sustained attention training and mental practice; Bowen, Hazelton, Pollock, & Lincoln, 2013). These methods seem to be chosen above others by experts worldwide to treat spatial neglect (Pen, Pitteri, Gillen & Ayyala, 2018), however there is currently no specific recommended rehabilitation method for the condition due to lack of high-quality clinical trials with significant positive results (Bowen et al., 2013).

The fast advancement of affordable, user-friendly and portable virtual reality (VR) technology means that it can be more easily applied in clinical settings (Castelvecchi, 2016). In fact, several VR interventions have been recently introduced for cognitive and/or physical impairments for various conditions, such as Parkinson’s disease (Dockx et al., 2016), Traumatic Brain Injury (TBI; Maggio et al., 2019) and stroke (Laver et al., 2017; Warland et al., 2019). VR enables a user to interact with a virtual environment, using either immersive (e.g. wearing VR headsets) or non-immersive technology (e.g. presents a virtual/computerized environment without being immersed within it). VR presents many advantages when compared to traditional rehabilitation including the ability to create rich sensory environments and replicate real-world situations within safe conditions (e.g. city; Cipresso et al., 2014; Ogourtsova et al., 2017), the ability to boost enjoyment, confidence and enthusiasm levels (Thornton et al., 2005; Pietrzak, Pullman & McGuire, 2014) and facilitate self-administration of rehabilitation while objectively monitoring therapy adherence remotely (Burdea, 2003; Threapleton, Drummond & Standen, 2016).

Telerehabilitation refers to the delivery of rehabilitation services remotely to patient’s homes using Information and Communication Technologies (ICT; Brennan, Mawson & Brownsell, 2009). Telerehabilitation can be delivered via real-time (synchronous) interactions between the patient and therapist using the telephone or video conferencing, or via ‘store-and-forward’ (asynchronous) methods where the patient’s progress is transmitted and reviewed by a therapist at a later time (Brennan et al., 2009; Richmond et al., 2017). Asynchronous telerehabilitation can be self-administered by patients in their homes, making them easier to implement by removing potential scheduling conflicts between patient and therapists (Hill & Breslin, 2016; Bini & Mahajan, 2017). Moreover, telerehabilitation can benefit patients living in remote locations and can be cost-effective when compared to one-to-one rehabilitation (Peretti et al., 2017). Importantly, randomised controlled trials exploring the effectiveness of telerehabilitation (mainly motor, e.g., balance training, mobility) have been successfully carried out (see Appleby et al., 2019 for a review). In fact, recent systematic reviews conclude that although the quality of the evidence (e.g., risk of bias, reporting quality) cannot yet show telerehabilitation is better than usual care, telerehabilitation (including VR) has comparable efficacy to face-to-face therapies (Sarfo, Ulasaverts, Opare-Sem & Ovbiagele, 2018; Appleby et al., 2019; Laver et al., 2020). More specifically, a number of studies have explored the feasibility of using home-based telerehabilitation techniques for stroke. For example, some of these studies found telerehabilitation techniques, such as physical rehabilitation using VR (Proffitt & Lange, 2015), upper extremity using a Home Care Activity Desk (HCAD; Huijgen et al., 2008), and game controllers to improve cognitive, upper extremity function and wellbeing (Burdea et al., 2020) and were positively rated and feasible to use in stroke survivors’ homes. However, there is little research on home-based VR rehabilitation for spatial neglect.

The development of VR rehabilitation for spatial neglect is still in its infancy. To date, studies in spatial neglect have explored both immersive and non-immersive VR including real-world tasks such as navigation and cooking (Ogourtsova et al., 2018; Tobler-Ammann et al., 2017) and visual search tasks (Yasuda et al., 2017; Cipresso et al., 2014). Fasotti and van Kessell (2013) reviewed several ‘proof-of-concept’ studies reporting positive effects of VR rehabilitation techniques for spatial neglect, such as simulated wheelchair navigation tasks (Webster et al., 2001), virtual street crossing (Katz et al., 2005), and searching and grasping tasks (Sedda et al., 2013). A randomised controlled trial by Kim et al., (2011), found a larger improvement in pre-and-post test scores on the Catherine Bergego Scale and star cancellation in neglect patients using interactive VR computer games, versus the control group using conventional neglect rehabilitation (e.g. puzzles, drawing, reading). More recently, Gammeri and colleagues (2020) state VR is an innovative and promising rehabilitation method for spatial neglect, which needs further exploration. Laver and colleagues (2020) suggest mixed-methods research is required in order to explore the feasibility and acceptability of these technologies in people’s homes. This would be essential in implementing telerehabilitation more widely, especially since experts report the intention to use VR based therapies for spatial neglect if they had optimal resources to facilitate its use (Pen et al., 2018).

Even though several VR therapies have been introduced for neurorehabilitation, not much is known about the views of stroke survivors, their carers or clinicians regarding their usability and acceptability (e.g., Threapelton, Drummond & Standen, 2016). The aim of this study was to explore the perspectives of end-users about non-immersive, self-administered VR telerehabilitation for spatial neglect. To the best of our knowledge, only one study has explored perspectives of using VR in spatial neglect but only investigated VR use for assessing neglect and only collected data from clinicians (Ogourtsova, Archambault & Lamontagne, 2019). Ogourtsova and colleagues used focus groups and interviews with clinicians and identified a series of barriers and facilitators including equipment (e.g., cost), client suitability (age), personal and institutional factors (e.g. familiarity with VR).

In the present mixed-methods study, we gathered perspectives on the use of self-administered VR telerehabilitation (asynchronous telerehabilitation) for spatial neglect from clinicians but also, importantly, from stroke survivors and their carers. Working and collaborating directly with end-users (e.g., strategies for patient-oriented research; SPOR; Canada’s Strategy for Patient-Oriented Research, 2011; Proffitt, Glegg, Levac, & Lange, 2019) our aims were to 1) identify any facilitators or barriers of use and 2) produce recommendations that may improve future development (Threapleton, Drummond & Standen, 2016) and, ultimately, enhance adherence, efficacy and implementation into clinical practise (Brouns et al., 2018). This study is an essential step in the development of future home-based VR telerehabilitation tools for spatial neglect, or indeed for other conditions (e.g., Lange et al., 2010), and a pre-cursor to conducting feasibility and efficacy studies in stroke survivors’ homes.

### 2.1 Participants

In line with similar studies (e.g. Ogourtsova, Archambault & Lamontagne, 2019; Lane et al., 2019; Niraj, Wright & Powell, 2018), our sample comprised of 16 participants: seven stroke survivors (SS; mean age = 67.1, *SD* = 6.8 years old, one female), three partners or carers of stroke survivors (C; mean age = 51, *SD* = 30.1 years old, one male) and six stroke clinicians (CL; one male, mean age = 44.7, SD = 12.1 years old). Demographic and clinical data of all participants are presented in Tables 1 and 2.

**Table 1.**
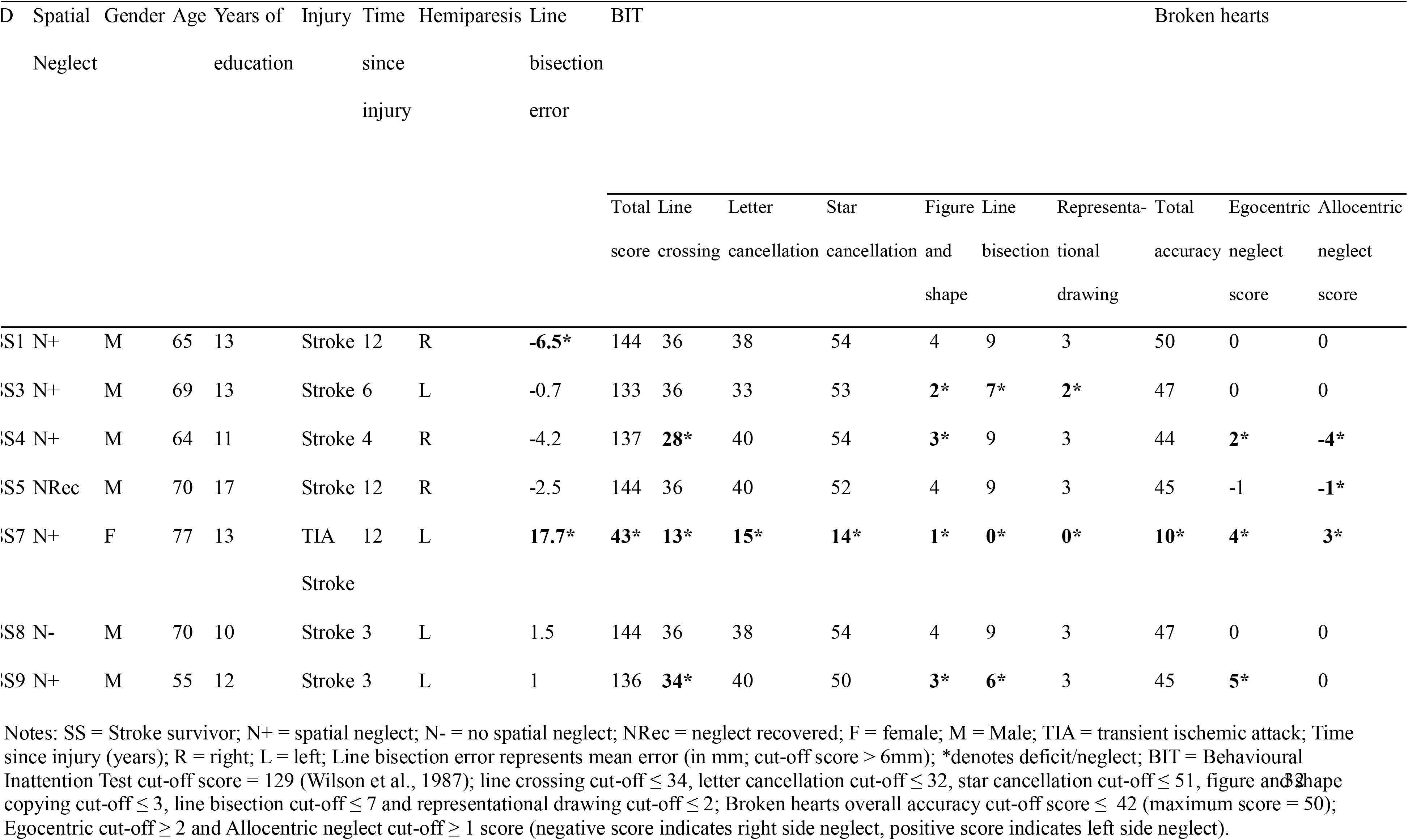
Demographic and clinical data for stroke survivors.

**Table 2.**
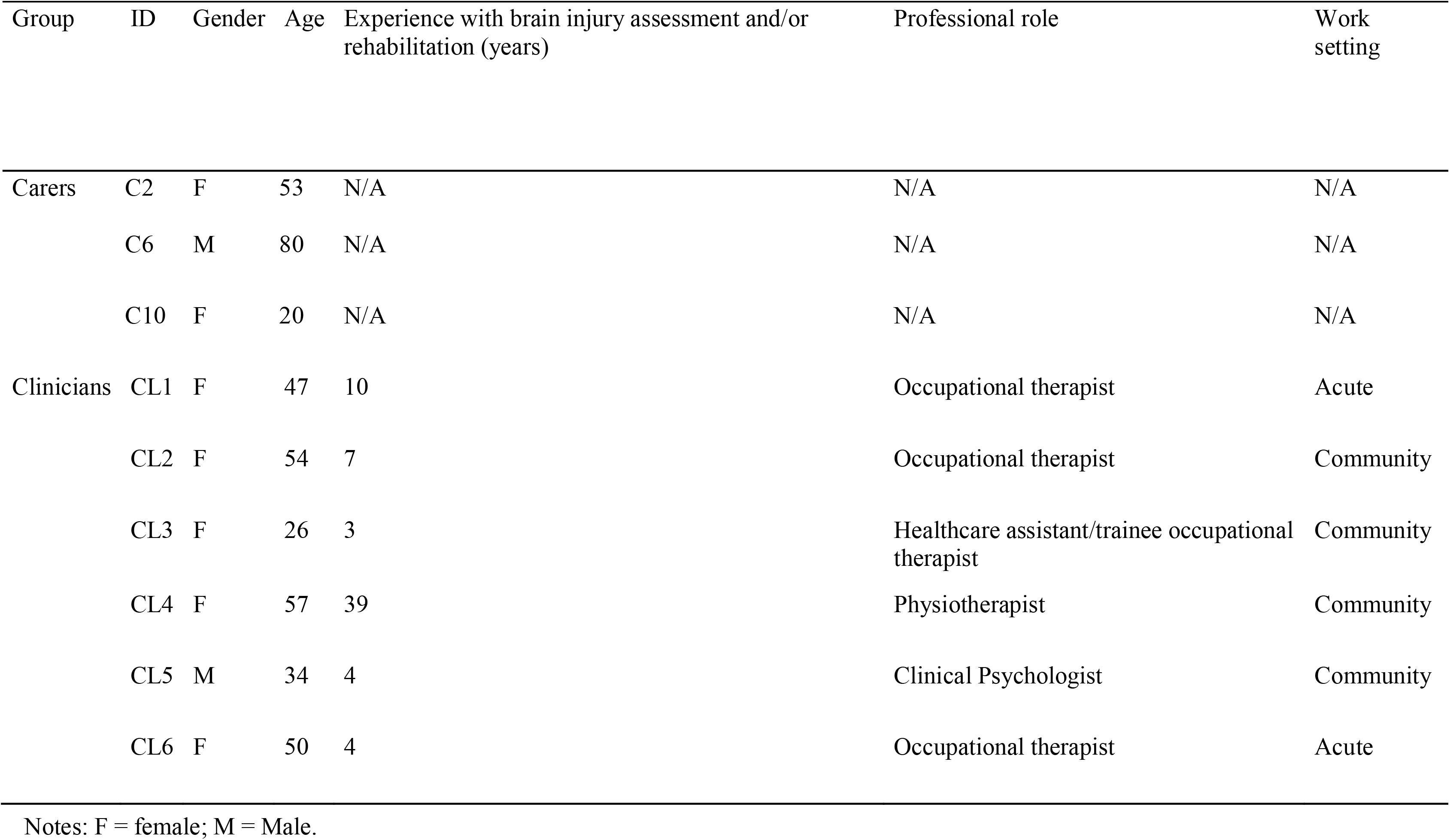
Carer and clinician demographic data.

All stroke survivors and carers were recruited face-to-face or by email/telephone from local community-led stroke support groups and social media. The inclusion criteria for eligible stroke survivors were: aged 18 years and over; history of stroke; fit to participate; able to give consent (as assessed by staff from recruiting site). Exclusion criteria were: signs of language impairment (which would severely affect production/comprehension); lack of capacity to consent. Carers were invited to take part if they were over the age of 18 and had experience caring for a stroke survivor. Two of the carers were partners of stroke survivors who also took part in the study. Time since stroke ranged between 3 and 12 years (mean = 7.4, *SD* = 4.4). Six stroke survivors self-reported experiencing spatial neglect in the past or present (e.g., could not see one-half of the television). None of the stroke survivors received any form of spatial neglect rehabilitation at the time of this study. Prior to focus groups, spatial neglect was formally assessed in a separate session at the participant’s home, using paper-and-pencil tests including Behavioural Inattention Test (BIT; Wilson, Cockburn & Halligan, 1987), line bisection task (Rossit et al., 2019) and the paper version of the Broken hearts test (Oxford Cognitive Screening; Demeyere et al., 2015). Five stroke survivors were classified as having chronic neglect at the time of the study (N+) as they were impaired in at least one of the spatial neglect tests (*n* = 3 left side neglect, *n* = 2 right side neglect). SS7 had the most severe neglect, presenting impairments on all tests, including a total score of 43 in the BIT. The remaining stroke survivors presented neglect on some but not all tests. One stroke survivor had recovered from spatial neglect (Nrec) and another did not show any symptoms in the tests used (N-; see Table 1 for neuropsychological test scores). The study was approved by the University of East Anglia Psychology ethics committee and all participants provided informed consent.

Six stroke clinicians (mean age = 44.7, *SD* = 12.1; Table 2) were recruited from local networks and took part in either individual interviews (N = 3) or a small focus group (N = 3). Experience working in brain injury (including working with patients with spatial neglect) ranged from 3 to 39 years (median = 5.5). Clinicians were only invited if they worked with brain injury survivors with spatial neglect at the time of the study.

### 2.2 Self-administered VR telerehabilitation

Telerehabilitation includes rehabilitation methods delivered using a variety of technologies, such as, but not limited to, therapeutic gaming technologies, mobile applications and/or virtual reality, in a number of settings (hospitals, community clinics, homes, institutional; Richmond et al., 2017). VirtualRehab is a telerehabilitation tool for motor rehabilitation using non-immersive VR and a motion-tracking sensor. Computerized spatial inattention grasping home-based therapy (c-SIGHT) is a non-immersive VR version of our home-based therapy for spatial neglect (grasping-to-lift rods at the centre; Rossit et al., 2019) which uses a motion-tracking sensor to track performance. The performance of participant can be monitored by a therapist remotely (live or at a later stage after the therapy has been completed).

During the first focus group (focus group 1), the stroke survivors and carers trialled three VirtualRehab ‘exergames’ (videogame-like exercise activities developed by Evolv Rehabilitation Technologies) including: ‘Boxing’ (sparring with a virtual boxing partner; Figure 1A), ‘Bullseyes and Barriers’ (hitting or avoiding targets; Figure 1B) and ‘In the Kitchen’ (search task in realistic kitchen layout; Figure 1C). During the second focus group (focus group 2), all participants trialled c-SIGHT (Figure 1D). Instructions were delivered on-screen and auditorily within the program throughout the focus groups and interviews, as well as verbally by the facilitator when required. VirtualRehab provided auditory (‘blings’, claps) and visual (stars, points) feedback while the participant controlled a virtual avatar. C-SIGHT also provided auditory (‘blings’) and visual feedback (frequency count) after completing an action correctly. C-SIGHT does not provide specific feedback about neglect stimuli as the user adjusts their performance based on the sensorimotor feedback from grasping-to-lift and balance rods (Rossit et al., 2019). Participants were told that when these tools are used in stroke survivors’ homes, the user’s performance would be monitored by therapists during or after use. The non-immersive VR was presented on a Samsung MD40B monitor (40-inch) attached to a laptop (HP 15.6 Omen) and used a low-cost and portable motion-tracking sensor (Microsoft Kinect) to measure body movements.

**Figure 1.**
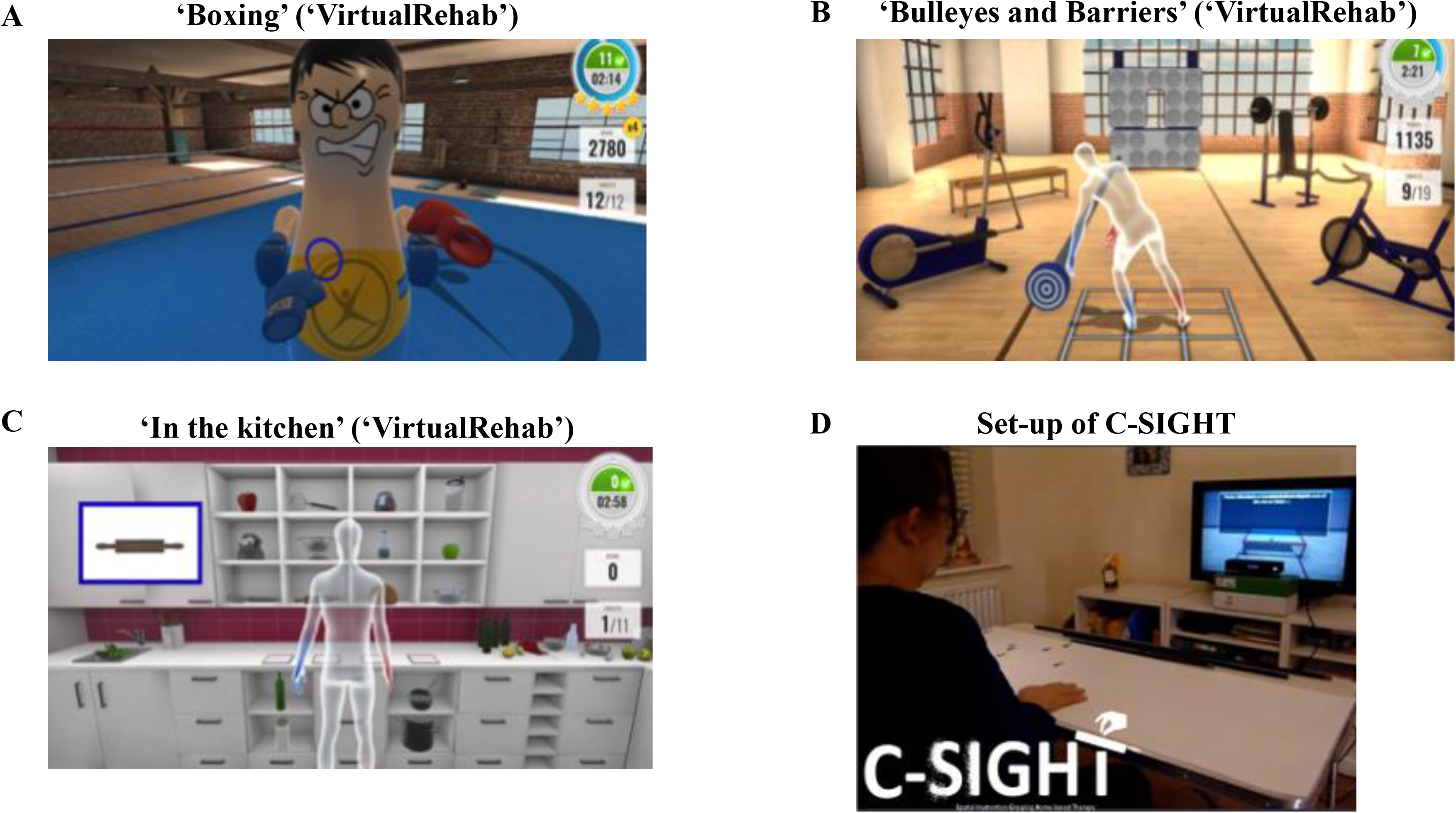
(A) display when using ‘Boxing’, (B) ‘Bulleyes and Barriers’, (C),’In the kitchen’ in ‘VirtualRehab’ (by Evolv Technologies) and (D) Set-up of C-SIGHT in a home setting.

### 2.3 Procedure

We carried out an explanatory sequential mixed-methods (Creswell, 2015) study using focus groups and semi-structured interviews combined with neuropsychological assessments and questionnaires (see flowchart in Figure 2). The study was primarily qualitative, with a quantitative component nested within the design (concurrent nested design; Clark & Creswell, 2008). Qualitative data was used in order to explore participant perspectives, and quantitative data was collected to enrich information on study participants, and aid interpretation of qualitative results (Morse, 1991).

Two separate focus groups were carried out with stroke survivors and carers (see Figure 2) in different dates at the University of East Anglia. Having separate focus groups was required to allow enough time for each participant to trial VirtualRehab and c-SIGHT separately and to reduce the overall length of each session. During focus group 1 we asked participants to trial VirtualRehab (*n* = 6 stroke survivors, *n* = 2 carers) and during focus group 2 participants trialled c-SIGHT (*n* = 6 stroke survivors, *n* = 3 carers). The same participants took part in both focus groups except SS8 who took part in focus group 1 only, and SS9 and C10 who completed focus group 2 only. Due to time constraints, clinicians took part in a single focus group/interview and trialled c-SIGHT only. However, clinicians were given information and shown pictures of VirtualRehab.

Before beginning focus group discussions, the moderator delivered a short presentation about the focus group guidelines (Vaugh, Schumm & Sinagub, 1996), study rationale and VR. Lead author and research associate working on the project (BSc, MSc) with experience of running a previous pilot focus group, conducted all focus groups and interviews. Following this, participants tried VirtualRehab (for the first time) and were encouraged to comment on this. Overall, each participant used the VR telerehabilitation for approximately 5 – 10 minutes in the same room as where the focus group took place. The focus group discussions followed 10 semi-structured questions and unscripted follow-up questions to further explore certain perspectives. Example of questions included: What did you find enjoyable in the virtual reality games? How did you find the feedback you received during the virtual reality games? What do you think about using this equipment and games at home or in clinic? (see supplementary materials for all questions). Individual interviews were carried out with three clinicians who could not attend the clinician focus group following the same procedure as the focus groups. Focus groups and interviews were audio recorded and transcribed by the lead author.

Following the focus group/interviews, to measure the usability of the VR telerehabilitation shown, all participants completed the System Usability Scale (which included statements such as ‘I thought the system was easy to use’; Brooke, 1996; Meldrum et al., 2012; Pei, Chen, Wong & Tseng, 2017). Other statements measured the intention to use the system, system integration, inconsistencies and confidence using the system. Scores on the System Usability Scale ranged from 0 (‘Worst Imaginable’) to 100 (‘Best imaginable’; Bangor, Kortum, & Miller, 2008; Bangor, Kortum & Miller, 2009). Moreover, participants also completed the ‘technology usage questionnaire’ (adapted from Vekiri & Chronaky, 2008) to investigate the frequency and variation of technology used by participants, including previous experience with VR. In addition, to explore the acceptance of using VR in their workplace (Ogourtsova et al., 2019), the clinician group completed the ‘Unified theory of acceptance and use of virtual reality questionnaire’ (Venkatesh, Davis & Davis, 2003). Finally, during the home visit for stroke survivors we collected information about home resources (WiFi, room size, TV size, TV model, table size).

**Figure 2.**
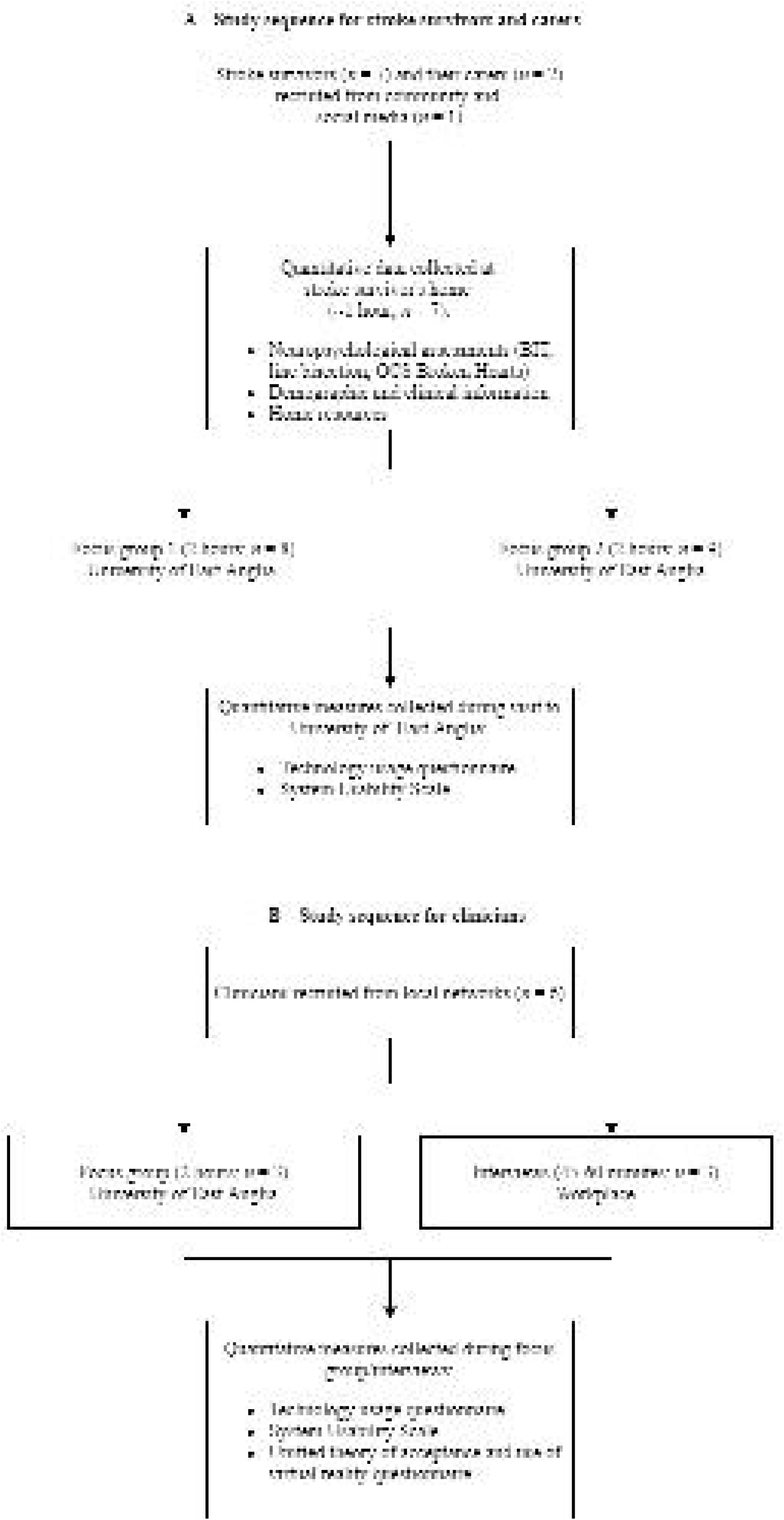
Flowchart of study for stroke survivors and carers (A) and clinicians (B).

### 2.4 Data analysis

Focus group and interview data was transcribed using Nvivo (QSR International’s NVivo version 12) by the first author (H.M). The data was analysed using a deductive thematic analysis with a semantic approach (Clarke & Braun, 2014). NVivo software was used to facilitate coding the data. Meaningful themes emerged across all focus group and interview data and were transposed to a coding grid to capture the facilitators and barriers to engagement with the VR telerehabilitation. Codes and themes were discussed by the research team to achieve consensus as to their meaning and/or overlap. A final round of analysis was then performed by author H.M until data saturation occurred across themes. Descriptive statistics were used to summarize quantitative data (e.g. frequency counts). All participants completed all required parts of the study and there were no drop-outs.

## 3. Results

### 3.1 Qualitative data

Data collected from focus groups and interviews revealed three major barriers and four major facilitators of using self-administered VR telerehabilitation for spatial neglect. Themes are described below and details of participant perspectives are shown in Table 3.

**Table 3.** Qualitative data. Details of stroke survivors (SS), carer (C) and clinician (Clin) perspectives for each theme.

| Theme (barrier or facilitator) | Participants' perspectives |
| --- | --- |
| Clarity of instructions (barrier) | SS9: As a stroke survivor, you need short instructions.<br>SS7: You don't want too long sentences<br>C2: I wondered if there were too many instructions though, if somebody was still really tired<br>Clin4: My concern would be people with attentional deficits, they would struggle a little bit<br>Clin1: Just a video of someone doing it themselves. |
| Level of experience with technology (barrier) | SS9: Different generations aren't used to using motion sensitive software.<br>SS5: I don't own a laptop. [VR rehabilitation is] designed by young people who are used to games<br>Clin1: My parents, who are in their 60s, 70s {...} would really struggle. It's fun {...} if you've got a young client.<br>Clin5: Younger patients might be more disposed to doing that.<br>Clin4: Even older people you can tell them and they can learn {...} Patients are quite used to technology now. |
| Equipment (barrier) | SS9: They might find it a bit of an intrusion {...} It's not so much it being on, as it just being physically there.<br>SS4: You'd be looking at it all the time.<br>SS3: You need to be careful because the CIA can get into them<br>SS5: If it becomes difficult to set up all the other benefits of the thing disappear.<br>SS8: What about the costing of this? Because not everybody can afford {this}<br>SS5: Expensive televisions or very expensive computers {...} you put people off<br>Clin2: Concerned about the amount of people who would have a TV {...} that big<br>Clin5: You just want to know in advance that they've got all the stuff [equipment] there. |
| Performance feedback (facilitator) | SS5: Virtual reality makes all the difference {...} Important to have a choice.<br>SS3: I think it's better with the computer, because it's as if there was somebody else you're doing it with {...} motivation to keep going<br>SS7: I found it encouraging<br>SS9: Some positive feedback in there would be hugely beneficial<br>C6: You wouldn't get any feedback without the computer.<br>C6: It would be interesting to see the progress she [SS7] makes.<br>Clin1: Compared to a therapist telling you what to do, it's a nice variation.<br>Clin5: The feedback very much comes from {...} the patient. |
| Engagement and enjoyment (facilitator) | SS3: If there's just a bit of fun involved, so it [would] definitely improve, increase the amount of time I spend on rehabilitation {...} they're exercises encouraging me to do exercise that I might not ordinarily do.<br>C2: Compete with each other. |
| Psychological benefits<br>(facilitator) | C10: Haven't got anyone standing over you, watching you do it, just take your time with it, make sure you're doing it properly and at your own pace {...} confidence thing, the independence as well, because you're doing it on your own.<br>SS5: I can see it being a significant advantage to people's psychological wellbeing.<br>SS5: We shouldn't underestimate just how significant of an advantage this would be to someone who had just had a stroke {...}<br>SS5: I was dependent on somebody, I had lost everything, and if this had come along and said, have a go at this and try it out, I would have given it a go and I would have, it would have made me feel a lot better about myself.<br>SS9: Hugely beneficial<br>Clin5: The patient has more, sort of ownership {...} this would be more motivating for a patient, because they're relying on their own feedback. |
| Meeting an unmet need<br>(facilitator) | SS5: Thinking back to how I felt immediately after the stroke, I would have certainly used this sort of thing {...} something you could do at home would certainly help.<br>SS8: It's easy {...} you don't have to go anywhere.<br>C6: The time you've got to spend getting ready and going to the gym and then coming home again you could actually do this.<br>SS9: As a tool for like, the early support discharge team, I think it would be really useful.<br>Clin1: This is a real way of measuring.<br>Clin2: There isn't anything else, often it's overlooked. |

#### Clarity of instructions (barrier)

All participants identified the length and format of instructions as important barriers of use in VR telerehabilitation. Lengthy instructions could be problematic for stroke survivors who experienced fatigue or cognitive impairments (“*My concern would be people with attentional deficits*” Clin4; Table 3). Instead of text alone, one clinician and stroke survivor suggested providing an option for users to have pictorial or video instructions.

#### Level of experience with technology (barrier)

Lack of experience or confidence with technology emerged as a potential barrier for two stroke survivors and one clinician. This was a particular concern as users might experience a technical problem which would worry them (“*different generations aren’t used to using motion sensitive software*” SS9). Similarly, two clinicians felt VR would be particularly desirable for younger clients, but one clinician noted that if implemented into clinical practise this technology would become more familiar and acceptable with older clients.

#### Equipment (barrier)

Some stroke survivors (3 out of 7) were concerned that the motion-tracking camera positioned above their television might pose a security risk and be an “*intrusion*” (SS9). In addition, one stroke survivor, carer and clinician felt that VR telerehabilitation would be less accepted and user-friendly if it was difficult to set-up (“*if it becomes difficult to set up all the other benefits {…} disappear*” SS5). Furthermore, four stroke survivors identified expensive equipment as a barrier of use, since not everyone would own or could purchase a laptop or television. Community and acute stroke clinicians (3 out of 6) were concerned that stroke survivors may not have adequate resources in their homes to accommodate the VR equipment (e.g. a television, space, Wi-Fi). Nevertheless, when asked directly about the equipment they trialled, all stroke survivors said they would be happy having the equipment in their homes.

#### Performance feedback (facilitator)

All participants believed that using VR telerehabilitation for spatial neglect would provide more feedback than conventional therapies. Stroke survivors and carers believed clear progress feedback (visual or auditory) would motivate the user and make rehabilitation more engaging. For example, SS3 found auditory prompts (e.g., cheers, claps) engaging by reassuring he was completing the therapy correctly (“*I like being cheered*”). However, one stroke survivor preferred discrete auditory prompts and feedback. Additionally, one carer thought it would be interesting to monitor their partner’s progress using the feedback from VR.

#### Engagement and enjoyment (facilitator)

One stroke survivor felt that ‘gamification’ of a therapy would increase their engagement, enjoyment and “*increase the amount of time I spend on rehabilitation*” (SS3). Two stroke survivors and one carer believed that adding a competition element to rehabilitation (either against the software, carer or other users) would increase their enjoyment.

#### Psychological benefits (facilitator)

Four stroke survivors shared the negative feelings they felt after their stroke (e.g. “*I was dependent on somebody, I had lost everything*” SS5, “a *lot of things in your life, your world have been taken away from you*” SS9). C10 thought self-administering rehabilitation would enable stroke survivors to go at their own pace. As a result, one stroke survivor, carer and clinician thought the user might feel more competence and “*ownership*” (Clin5) over their rehabilitation since they could carry it out independently. Moreover, two stroke survivors and one carer thought this would increase positive feelings, such as confidence and independence (e.g. “*made me feel a lot better about myself*” SS5).

#### Meeting an unmet need (facilitator)

When asked their opinion on using VR for rehabilitation, two stroke survivors thought it was an “*excellent idea*” (SS3, SS4) and *“very good idea*” (SS5). Stroke survivors (2 out of 7) and carers (2 out of 3) believed a self-administered VR telerehabilitation for spatial neglect would be more convenient and accessible. One stroke survivor noted that home-based VR telerehabilitation would be especially beneficial for those who had mobility issues. Moreover, all clinicians recognised the unmet need for spatial neglect rehabilitation. In addition, they also recognized the benefits of developing a self-administered VR telerehabilitation for spatial neglect with the capabilities of objectively measuring adherence remotely.

### 3.2 Quantitative data

#### 3.2.1. System Usability Scale (SUS)

All participants gave an adequate SUS score regarding the VR telerehabilitation tools for spatial neglect: stroke survivors (62; ok), carers (74.2; good) and clinicians (67.5; ok). This indicates that the self-administered VR telerehabilitation was acceptable for all participants (Bangor, Kortum, & Miller, 2008; Bangor, Kortum & Miller, 2009).

#### 3.2.2. Technology-usage questionnaire

All but two stroke survivors (SS7, due to severe neglect symptoms) reported owning a mobile phone or smartphone. Clinicians and carers reported using a laptop/computer daily or 2-4 times a week, whereas stroke survivors’ computer usage varied from daily to rarely. Only one stroke survivor played computer games daily. All stroke survivors and the majority of clinicians had never tried VR before the present study (see Figure 3A). More than 67% of stroke survivors and carers were extremely interested in using VR in their rehabilitation and 66% of clinicians were very interested (see Figure 3B). All participants were mostly confident that they could use VR for rehabilitation (see Figure 3C).

**Figure 3.**
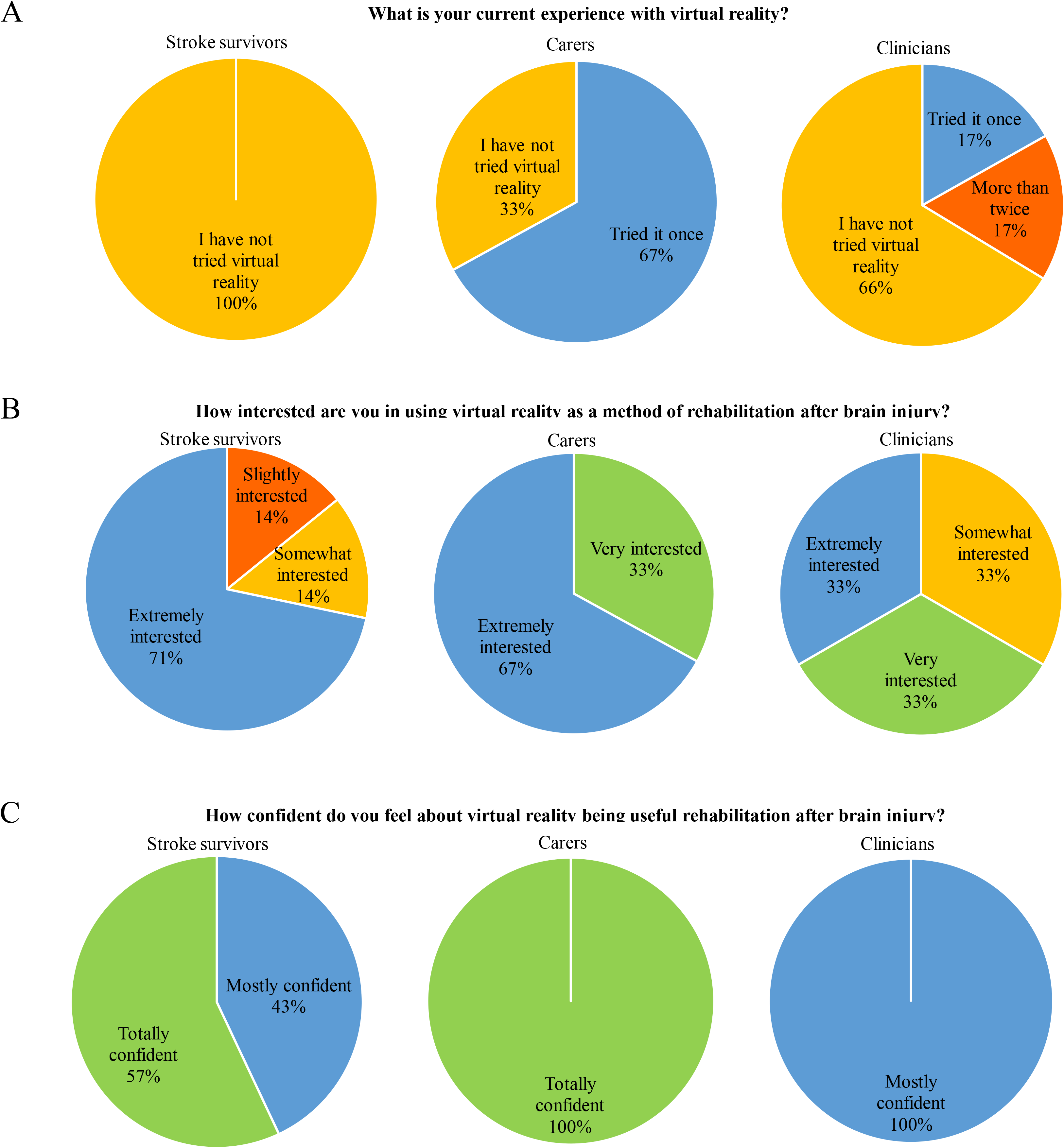
(A) What is your current experience with Virtual Reality?, (B) How interested are you in using virtual reality as a method of rehabilitation?, (C) How confident do you feel about virtual reality being a useful rehabilitation after brain injury?

#### 3.2.3. ‘Unified theory of acceptance and use of virtual reality ‘ for spatial neglect questionnaire

All clinicians (except one) agreed that VR rehabilitation could improve their work performance and effort (see Q1-5 in Fig.4). Clinician’s reported mixed views on whether members of their organization would support the use of VR rehabilitation (see Q6-9 in Fig.4). The majority of clinicians (80%) reported having adequate resources and knowledge to use VR rehabilitation in clinical practice (see Q10-16 in Fig.4). Finally, most clinicians (67%) agreed that they intend to use VR rehabilitation if available in the next year (see Q17 in Fig.4). Notably, one clinician noted on questionnaire that they would plan to use VR rehabilitation in the next 12 months “*if I had the support from management and time to use*”.

**Figure 4.**
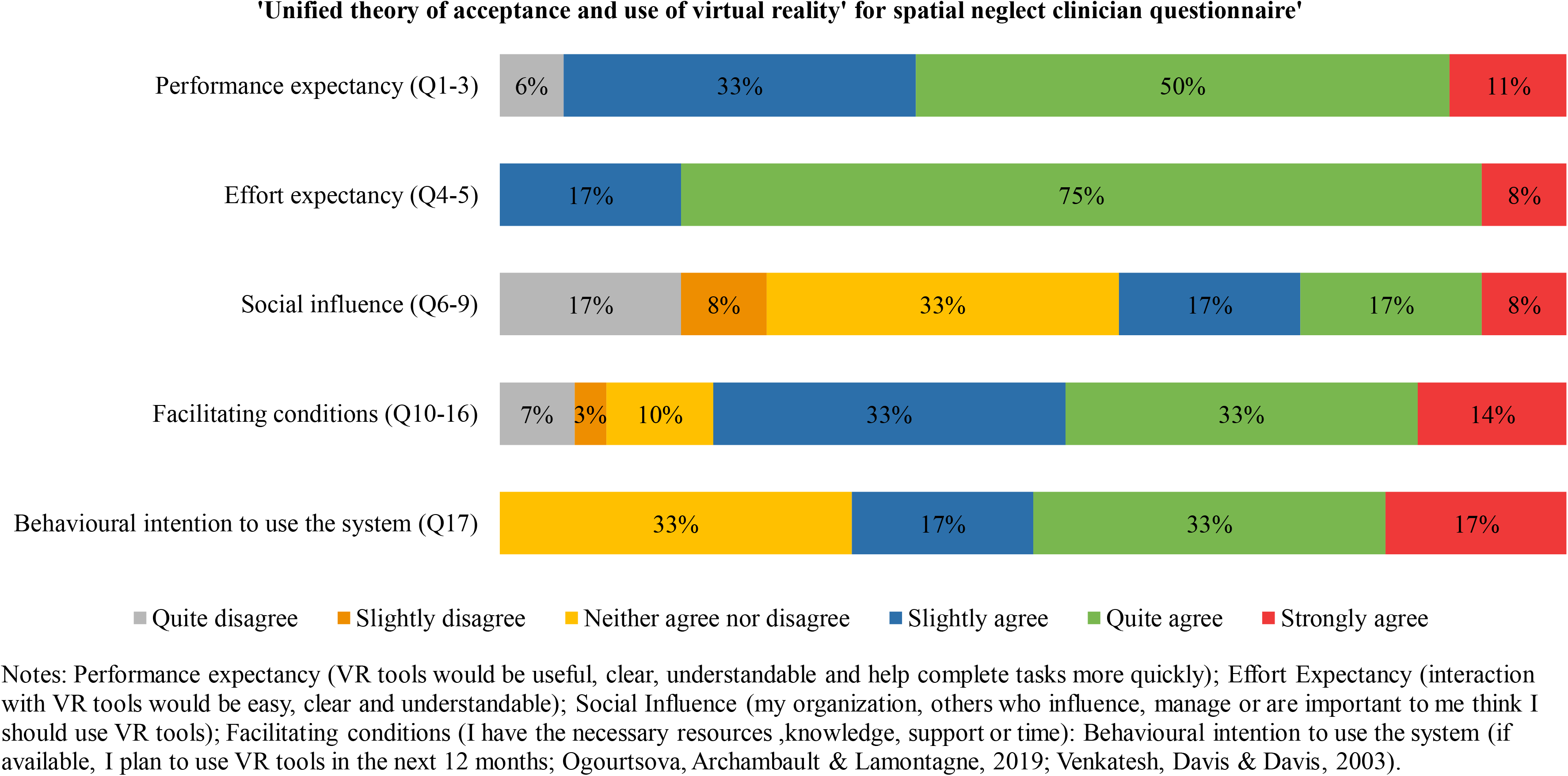
‘Unified theory of acceptance and use of virtual reality’ for spatial neglect clinician questionnaire

##### Home resources

All stroke survivors had a television larger than 32 inches and Wi-Fi which would enable them to run VR telerehabilitation from their homes. Moreover, all stroke survivors had an average of 5.3ft in front of their television which would allow them to use the motion tracking sensor to record body movements.

## 3. Discussion

To the best of our knowledge, this is the first study to explore perspectives on self-administered VR telerehabilitation for spatial neglect with stroke survivors, carers and clinicians. We identified three barriers and four facilitators of using self-administered VR rehabilitation for spatial neglect which can be considered in future to increase its acceptability and usability among users. Importantly, this mixed-methods design revealed that end-users were accepting and willing to use a self-administered VR telerehabilitation for spatial neglect. Both qualitative (barriers and facilitators) and quantitative data (e.g., resources in stroke survivor’s homes, SUS scores) not only inform the development of self-administered VR telerehabilitation for spatial neglect, but also the design and feasibility of running a future home-based pilot study to explore the usability of these tools in an ecologically valid setting.

Focus groups and interviews identified the need for short instructions and use of multiple formats (e.g. pictures, large font; similar to Wentink et al., 2019). This is important since those using self-administered VR telerehabilitation for spatial neglect (e.g. c-SIGHT) may experience difficulty with functional reading (associated with increased neglect severity; Galletta et al., 2014). Additionally, considering around 30% of stroke survivors are affected by aphasia (impaired language and communication function; Dickey et al., 2010) the provision of pictorial and/or auditory instructions would mean these stroke survivors would not be excluded from using self-administered VR telerehabilitation for spatial neglect.

Interestingly, it seemed clinicians and stroke survivors had an expectation that those with little experience with technology or of older age are not interested in using VR (e.g. Farrow & Reid, 2004; Ogourtsova et al., 2019; Wentink et al., 2019). Indeed, a stroke survivor with less technological experience did express a concern that VR telerehabilitation would not be accepted if it was difficult to set-up. However, participants (many reporting little technological experience) did not acknowledge this as a barrier. For example, a stroke survivor (SS3) and carer (C2) felt confident about setting up the equipment since they had previous experience (e.g. Wii Fit). In fact, participants (notably clinicians) were very interested and felt confident in using VR telerehabilitation and rated its usability as acceptable, regardless of their technology usage/experience.

Participants valued the features facilitated by VR equipment (e.g. remote monitoring of adherence, feedback) and would be willing to use it in their homes. However, some stroke survivors believed user acceptance could be affected by the risk of security breaches and the presence of the motion tracking camera in their home (e.g. aesthetics; Threapleton, Drummond & Standen, 2016). In this study, our sample had resources in their homes to support self-administered VR, nevertheless it is important to consider this since the other users’ homes may not have these resources due to cost (e.g. large TV). Therefore, changes may have to be made (e.g. move furniture/items) to accommodate the equipment, which may not be acceptable or feasible to the user (e.g. if they use assistive equipment; Threapleton, Drummond & Standen, 2016; Mountain, Wilson & Eccleston, 2010).

Previous findings have found clinicians report direct feedback from VR to be beneficial in motor learning principles and increasing therapy usage (Schmid, Glassel & Schuster-Amft, 2016; Finley & Combs, 2013). Similarly, our study found feedback provided by self-administered VR telerehabilitation (e.g. auditory prompts, visual progress) was seen as a major facilitator of use (e.g. Burdea, 2003). A stroke survivor felt feedback offered by self-administered VR telerehabilitation would increase the enjoyment and engagement he felt while carrying out “*ordinarily boring*” (SS3) exercises/tasks. Among feelings of engagement and motivation, participants believed users might feel an increase in competence, confidence, independence and ownership from using self-administered VR telerehabilitation. Farrow and Reid (2004) found similar results with stroke survivors who felt VR enabled them to engage in activities they could no longer do, increasing feelings of competence, control, and positive feeling of self (Farrow & Reid, 2004; Pallesen et al., 2018). Considering one-third of stroke survivors report experiencing symptoms of post-stroke depression (Stroke Association, 2018; Hackett et al., 2014), this is an important perceived benefit of using VR telerehabilitation.

Stroke clinicians recognised that there was a need for a rehabilitation for spatial neglect, especially one with remote monitoring/accessibility to user performance facilitated by VR (Burdea, 2003; Ogourtsova et al., 2019). Whereas, stroke survivors and carers believed the self-administered aspect of VR telerehabilitation would increase the accessibility of rehabilitation for spatial neglect. Stroke survivors in this study believed offering self-administered VR telerehabilitation to stroke survivors after discharge could help those with decreased mobility and increase their psychological wellbeing. This is an important perceived benefit since around 45% of stroke survivors report feeling abandoned after discharge (Stroke Association, 2018). Reflecting on a carer’s comment on convenience, offering self-administered VR telerehabilitation for spatial neglect at home could reduce the cost and time for carers (Tindall & Huebner, 2009).

We collected perspectives from a multidisciplinary group of stroke clinicians (occupational therapists, healthcare assistant, physiotherapist and clinical psychologist). Previous research has suggested the use of a VR rehabilitation is influenced by the clinician’s current technology experience (Burdea, 2003). Although these clinicians reported a low to moderate technology usage, they were open and positive about using VR telerehabilitation and believed it could improve their performance during clinical practice (Ogoutsova et al., 2019). While most clinicians had adequate resources to support VR telerehabilitation usage, clinicians working in an acute setting (e.g. hospital) anticipated more difficulty since there were limited resources to facilitate VR use (e.g. lack of working computers, space, television screens). Facilitating conditions such as these are a powerful predictor of a clinician’s intention to use a new technology (Liu et al., 2015) and therefore are an important factor to consider when planning implementation of VR telerehabilitation into clinical practice. Perspectives collected from this multidisciplinary group are promising indications that stroke clinicians within different settings would be willing to implement self-administered VR telerehabilitation for spatial neglect with their patients.

Some studies have explored clinician perspectives on VR rehabilitation for upper limb training (Schmid, Glassel & Schuster-Amft, 2016), exergames (Nguyen et al., 2019) and immersive games (Farrow & Reid, 2004). However, few (e.g. Pallesen et al., 2018) benefit from exploring perspectives from both clinicians and stroke survivors. Moreover, despite the emergence of using VR rehabilitation interventions for spatial neglect (e.g. Ogourtsova et al., 2018; Tobler-Ammann et al., 2017; Yasuda et al., 2017; Cipresso et al., 2014) few studies qualitatively explore perspectives of end-users. The current study used mixed-methods which offered a holistic approach (Klinke et al., 2016) to explore stroke survivor, carer and clinician’s perspectives, whilst objectively measuring factors (e.g. neglect, stroke severity, technology usage, resources) which might influence perspectives.

However, the current study has limitations. Due to time constraints only one focus group was held with clinicians. This could be considered a limitation since one large focus group with all six clinicians may have facilitated deeper exploration of ideas and opinions with others (Folch-Lyon & Trost, 1981).

Despite our relatively small sample size (albeit in line with sample sizes of similar studies; Lane et al., 2019; Ogourtsova et al., 2019; Pallesen et al., 2018; Niraji, Wright & Powell, 2018), we were able to collect the personal perspectives of various end-users (stroke survivors with and without spatial neglect, carers of different ages and multidisciplinary stroke clinicians). Nevertheless, since we used convenience sampling to recruit community-dwelling stroke survivors from stroke community groups, there may have been a sampling bias in the data. That is, the views obtained here may not fully represent the opinions of all stroke survivors, especially those who do not attend stroke community groups. Future research should recruit participants from stroke services and those in different stages of their recovery. Lastly, although including clinicians from a variety of backgrounds, with different level of experience and from different settings may have been beneficial to collect a range of perspectives, it made it more difficult to interpret questionnaire responses (e.g., resources varied depending on the work environment).

Our findings are both original (e.g. first to explore perspectives on self-administered VR telerehabilitation for spatial neglect) and consistent with previous studies (e.g. Ogourtsova et al., 2019). Feedback from this study will be invaluable in the development of novel rehabilitation tools for spatial neglect and future trials. Involving clinicians, patients and carers in a collaborative design process is not only recommended for the development of VR rehabilitation (Proffitt et al., 2019), but also ensures the user’s experience and priorities are incorporated (Santana et al., 2020). This in turn improves and informs tool development to tailor it to specific populations, compared to using ‘off-the-shelf tools (Lange et al., 2010). This study demonstrates the usefulness of involving patients/carers (i.e. strategies for patient-oriented research; SPOR; Canada’s Strategy for Patient-Oriented Research, 2011) early on in the development of such tools. Future studies should carry out a usability study to test the equipment and set-up of self-administered VR telerehabilitation for spatial neglect in stroke survivor’s homes (e.g. Warland et al., 2019). This would produce both ecologically valid and constructive feedback on the feasibility and acceptability of using self-administered VR telerehabilitation in user’s homes. Additionally, there is a need for more qualitative and mixed-methods studies exploring stroke survivor, carer and clinician perspectives on both non-VR and VR rehabilitation techniques for spatial neglect. Identifying factors which motivate and engage stroke survivors with spatial neglect provides us with a better understanding on how to increase therapy adherence and enjoyment.

Based on the perspectives from end-users in this study, we have produced some practical recommendations for future development of self-administered VR telerehabilitation for spatial neglect. To address concerns regarding user instructions we recommend using short text presented gradually and providing a choice of format for users (e.g. pictorial, written and/or auditory). Secondly, all efforts should be made to reduce the volume, complexity of equipment used (such as, using existing equipment in participant’s homes; e.g. television). Concerns could be reduced by labelling equipment (including cables), providing a clear, comprehensive demonstration of set-up and video or step-by-step instructions (with pictures). Embedding built in support (e.g. ‘help’ button, contact number) and online forum for users to ask each other questions are also possible steps to take. Thirdly, security concerns can be reduced by ensuring transparency of the use of each piece of equipment and recommending users switch off motion sensor cameras when not in use. Finally, the user should be provided with a choice to personalise their telerehabilitation when possible, such as providing a choice of visual or auditory feedback and music.

## Conclusion

This mixed-methods study identified that the acceptability of self-administered telerehabilitation was determined by the length and format of instructions. Additional barriers were the potential lack and cost of available resources for stroke survivors and acute setting stroke clinicians, which would facilitate usage of self-administered VR telerehabilitation for spatial neglect. Overall, stroke survivors, carers and clinicians were accepting, interested and confident about using self-administered VR telerehabilitation regardless of their level of technology experience. Potential psychological benefits were identified, such as an increase in independence and confidence, which were associated with motivating and engaging feedback provided by VR.

Future research on VR rehabilitation for spatial neglect should incorporate end-user perspectives to improve the acceptability and engagement of interventions. Perspectives collected using this mixed-methods design with three groups of end-users has enabled us to produce practical recommendations for future development of VR telerehabilitation post-stroke. It is hoped these recommendations may improve future development (Threapleton, Drummond & Standen, 2016) and increase user enjoyment and engagement with VR telerehabilitation.

Involving end-users in the early stages and throughout development of VR telerehabilitation tools informs the planning and design of more cost-efficient and effective research studies. The next steps of this research will be to conduct studies exploring the usability and feasibility of self-administered VR rehabilitation for spatial neglect in stroke survivor’s homes.

## Data Availability

N/A

## Acknowledgements

We would like to thank all the stroke survivors, carers and clinicians for their useful comments and kind cooperation during this study. We also thank the reviewers for their valuable comments and suggestions. This research was supported by the University of East Anglia Innovation Proof-of-Concept Fund 2017-2018. C-SIGHT developments were also funded by a grant to SR and VP from the National Institute for Health Research (NIHR) Brain Injury MedTech Co-operative based at Cambridge University Hospital NHS Foundation Trust and University of Cambridge. HM is currently funded by a grant from The Stroke Association to SR, HM and VP.

